# Assessing the repurposing potential of disease-modifying antirheumatic drug targets to reduce Alzheimer’s disease risk: a pQTL-based Mendelian randomization and colocalization analysis

**DOI:** 10.1101/2025.08.01.25332834

**Authors:** Christina N. Kushnir, Victoria Taylor-Bateman, Neil M. Davies, Emma L. Anderson

## Abstract

**Background:** Current literature has implicated systemic inflammation in the pathogenesis of Alzheimer’s disease (AD). However, the viability of anti-inflammatory drug targets as repurposed treatment candidates for AD remains unclear. We utilized two-sample Mendelian randomization (MR) and colocalization analysis to investigate whether disease-modifying antirheumatic drug (DMARD) targets reduce AD risk.

**Methods:** We investigated 9 DMARD targets, using blood protein quantitative trait loci (pQTLs) from the UK Biobank Pharma Proteomics Project (*n* = 54,219). The associations of these variants and AD were extracted from the International Genomics of Alzheimer’s Project (*n*cases = 21,982, *n*controls = 41,944). We used two-sample MR analyses to estimate the effects of perturbing DMARD targets on AD risk. We also examined the colocalization of target pQTLs and AD signals in the *cis*-regions of target-encoding genes.

**Results:** Our MR estimates suggested that a 1 SD increase in plasma protein concentration of the etanercept target, *FCGR3B*, increased the risk of AD by 10% (OR 1.10; 95% CI [1.02, 1.19]; *p* = 0.01). We found little evidence that the remaining DMARD targets reduced AD risk. Further colocalization analysis indicated no evidence of colocalization between encoding genes of DMARD targets and AD, including *FCGR3B*. Though, there was suggestive evidence of a non-colocalized association between *TNF* and AD on chromosome 6 via distinct causal variants (H_3_ = 46.5%).

**Conclusions:** Our findings suggest that the majority of analyzed DMARD targets were unlikely to reduce AD risk, except for *FCGR3B*. Future research in AD drug targeting and repurposing should examine the therapeutic viability of drugs which lower *FCGR3B* protein levels in blood plasma.

## Background

Over 50 million people worldwide suffer from dementia, with this number expected to increase nearly threefold by the mid-21st century [1]. Alzheimer’s disease (AD), the most common cause of dementia and a leading cause of death in the United States and the United Kingdom, imposes both immense pain on patients and their loved ones as well as a significant socioeconomic burden on healthcare systems, making it imperative to find treatment candidates that do not simply address quality of life but, more urgently, target pathological mechanisms that may prevent or halt disease progression [2, 3].

In recent years, chronic and systemic inflammation has been recognized as a possible modifiable risk factor for AD. Elevated concentrations of cytokines in the central nervous system (CNS) lead to neuronal toxicity and tissue damage which facilitate the susceptibility and development of AD [4, 5]. Additional dysregulation of key pro-inflammatory mediators, such as tumor necrosis factor (TNF-α) and interleukin-6 (IL-6), has been connected to hallmark molecular mechanisms contributing to the pathogenesis of AD [4, 5, 6, 7, 8]. Furthermore, genome-wide association studies (GWAS) have substantiated the role of immune function and inflammation in AD. The largest and most recent AD GWAS to date, conducted by Bellenguez et al. (2022), identified multiple genome-wide significant loci near genes involved in innate immunity and microglial function, such as *TREM2, PLCG2*, and *ABI3*, as well as loci near *IL-34, TNIP1*, and *GRN*, all of which have roles in immune regulation or inflammatory signaling [9]. These findings provide converging genetic evidence that dysregulated immune responses may contribute to AD pathogenesis, highlighting inflammatory pathways as promising therapeutic targets.

Disease-modifying antirheumatic drugs (DMARDs), a class of anti-inflammatory medications that modulate both cytokine activity and other pro-inflammatory targets, present as potentially viable drug repurposing candidates for AD prevention and treatment. Importantly, the disease-modifying component of DMARDs makes them encouraging therapeutic candidates to counter the progressive neurodegeneration that characterizes AD. However, observational studies examining the association between prescribed DMARD usage and the risk of AD have yielded mixed and contradictory results [10, 11, 12, 13, 14, 15]. Furthermore, such studies investigate sample populations already impacted by autoimmune and/or rheumatic diseases, making confounding by indication highly likely. While randomized control trials (RCTs) may help clarify the existing research, only two RCTs have investigated the casual effects of DMARDs on the risk of AD, both with small sample sizes (*n* = 41, *n* = 15) [16, 17]. These RCTs examined the use of etanercept, a commonly prescribed DMARD, over the course of 6 months with results from both trials suggesting positive trends in favor of a protective effect on AD risk. However, the lack of statistical power limits inference from these studies.

Mendelian randomization (MR) is a gene-based instrumental variable approach that can help to minimize confounding and reverse causation—which are pervasive in observational studies— without the additional logistic and financial constraints of clinical trials [18]. Due to the randomized nature of genetic inheritance, MR can assess the causal effects of risk factors via instrumentation of genetic variants [19]. Additionally, MR can be specifically employed to study the effects of perturbing drug targets [19]. Drug target MR estimates the effects of changing protein levels or expression under the rationale that these traits are likely to mimic the consequences of perturbing these proteins by drugs known to target these mechanisms. The genetic variants are selected based on their association with the drug targets of interest (Fig. 1). Colocalization often accompanies MR studies as a form of sensitivity analysis to reduce the threat that linkage disequilibrium (LD) poses to the assumptions of MR. As neither MR nor colocalization are alone sufficient in establishing causality, the integration of both methods may provide strength to studies in statistical genetics and drug repurposing.

**Fig. 1.**
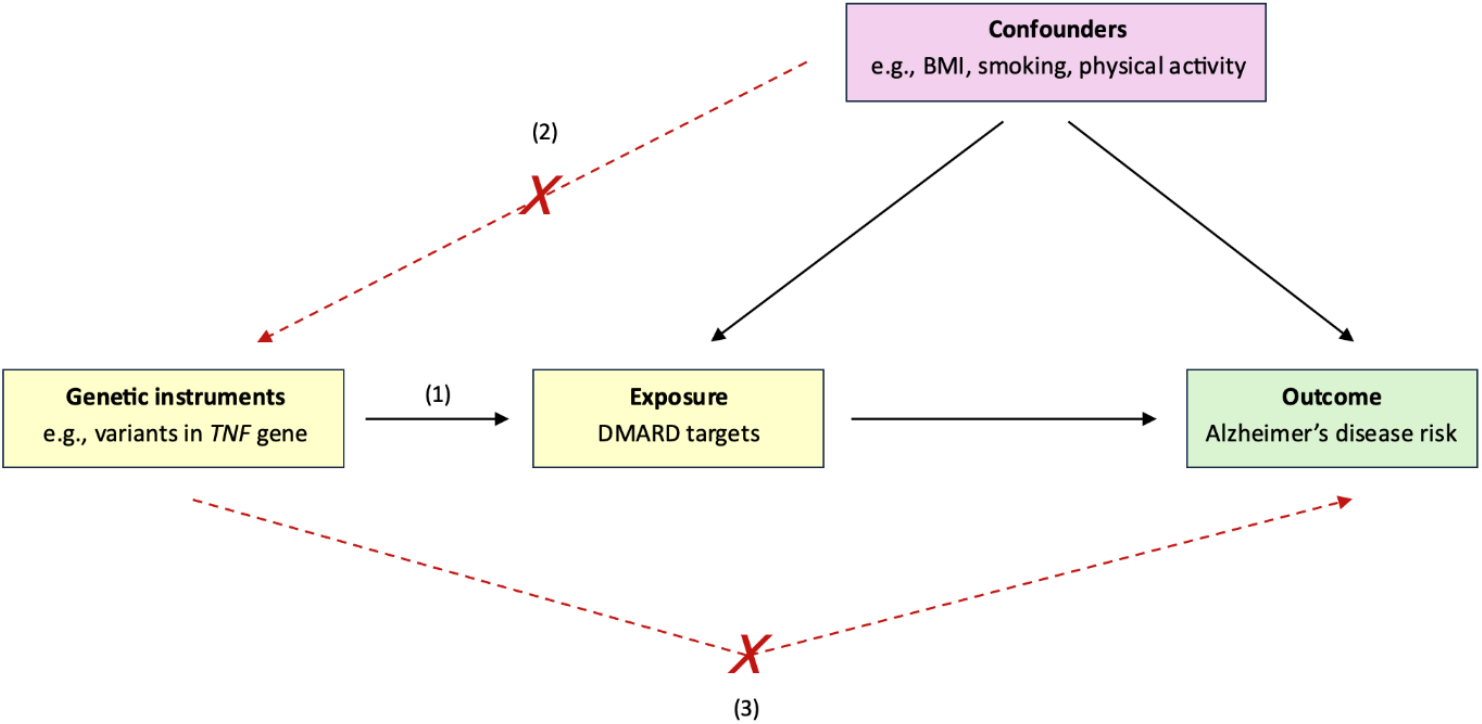
Diagrammatic illustration of the study. MR assumes that the genetic variant (1) is robustly associated with the exposure variable (relevance); (2) has no shared confounders with the outcome (independence); and (3) affects the outcome only through the exposure (exclusion restriction) [20]. Abbreviations: DMARD, disease-modifying antirheumatic drug; *TNF*, tumor necrosis factor; BMI, body mass index

Here, we assess whether select DMARD targets are likely to be efficacious therapeutic targets for reducing AD risk, using two-sample MR and colocalization.

## Methods

### Selection of DMARD targets

11 DMARDs, totaling 33 target genes, were initially selected for analysis. The rationale for inclusion was based on findings from the current literature regarding associations between DMARD usage and incidence rates of dementia or AD [10, 11, 12, 13, 14, 15]. The drugs identified were methotrexate, sulfasalazine, hydroxychloroquine, leflunomide, adalimumab, etanercept, infliximab, certolizumab, golimumab, tocilizumab, and tofacitinib. Methotrexate and tofacitinib had no available pQTL data and were therefore excluded from analysis. In later stages, the pQTL data for the leflunomide target, *DHODH*, did not meet the required significance threshold; as a result, leflunomide was also excluded from analysis. DMARDs and their corresponding encoding gene targets were identified using the DrugBank database (https://go.drugbank.com) and are listed in Table 1. Note, this table does not encapsulate all targets of the respective DMARDs but rather the targets with available pQTL data. Additionally, some targets, such as *TNF*, are shared across multiple drugs in our analysis.

**Table 1:**
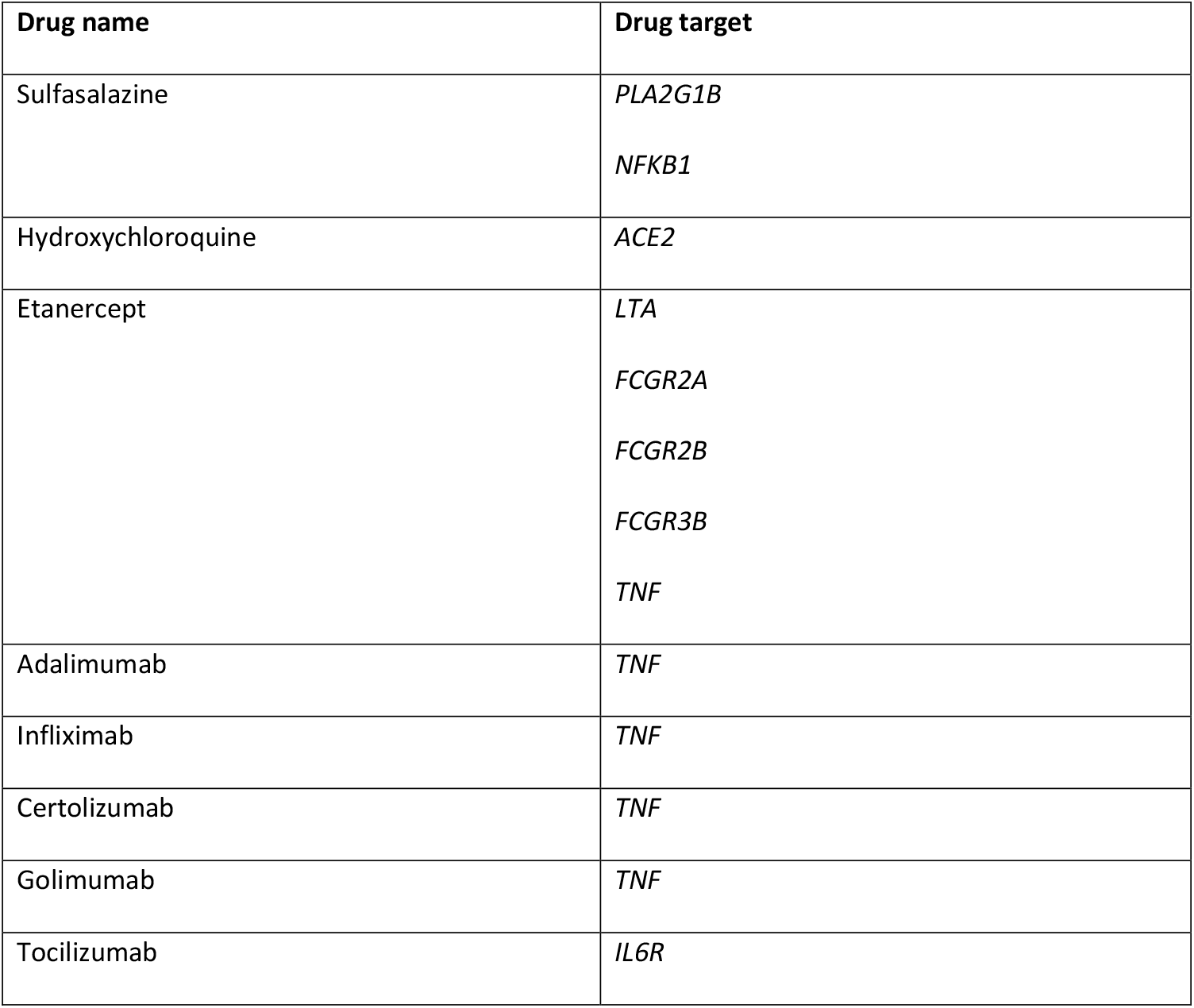
Selected DMARDs and their analyzed targets.

### pQTL instrument selection

Exposure data for each selected DMARD target were sourced from a large-scale proteogenomic analysis as part of the UK Biobank Pharma Proteomics Project (UKB-PPP, *n* = 54,219), which provides detailed mapping of 2,923 proteins and their associations with genetic variants within blood plasma [21]. We searched for *cis*-acting single nucleotide polymorphisms (SNPs) known to be associated with circulating protein levels (pQTLs) of encoded DMARD targets (Fig. 2). Lead pQTLs were identified to proxy the effects of select DMARD target perturbations, based on independence and strength of association (meeting the genome-wide significance threshold of *p* < 5 × 10^−8^). The following selection criteria were applied for instrumentation: 1) pQTLs within ± 1000 kb of the gene region (*cis*-acting); and 2) a genome-wide significance threshold of *p* < 5 × 10^−8^).

**Fig. 2.**
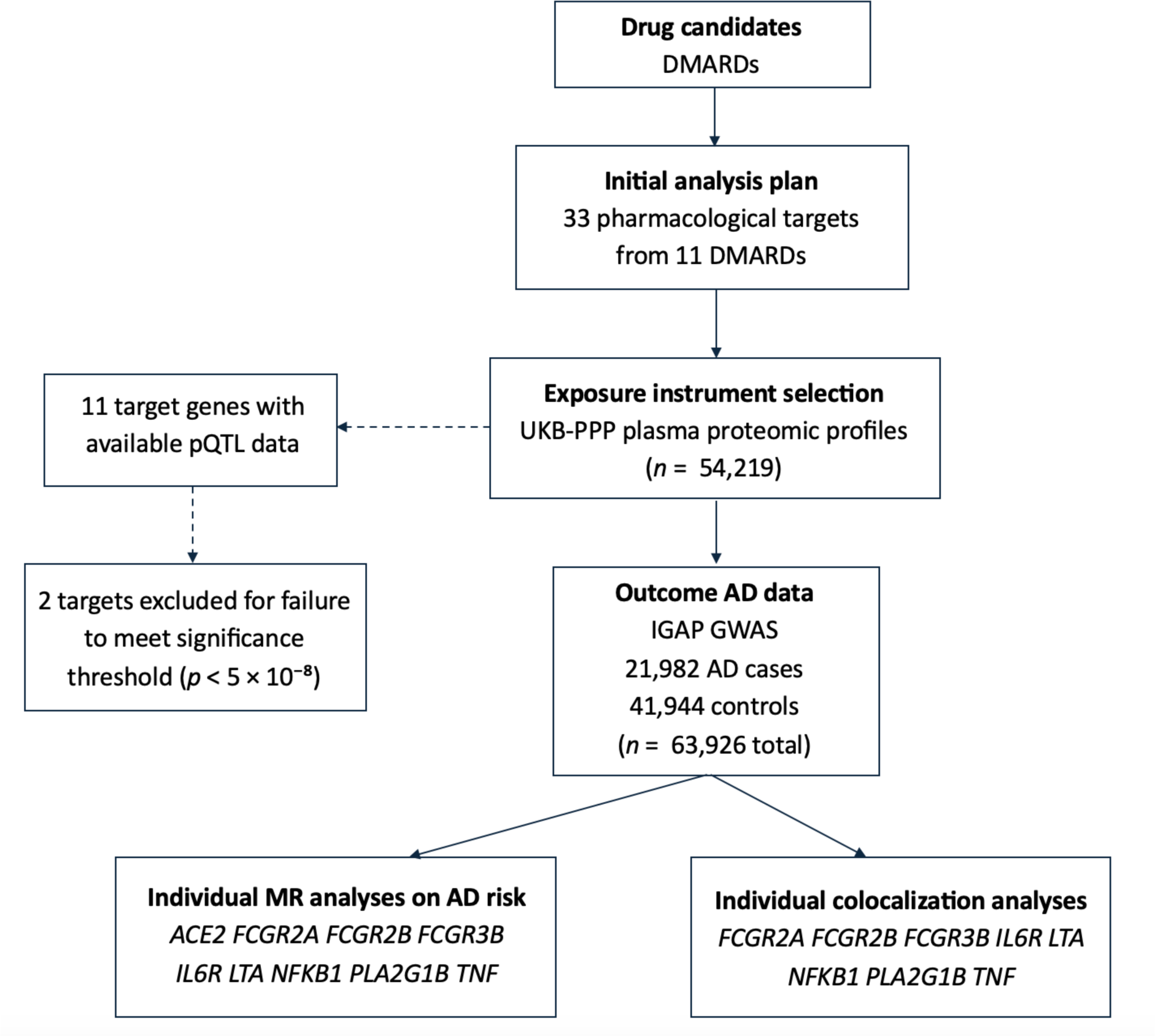
Flow diagram of the study design. Abbreviations: DMARDs, disease-modifying antirheumatic drugs; UKB-PPP, UK Biobank Pharma Proteomics Project; pQTL, protein quantitative trait loci; AD, Alzheimer’s disease; IGAP, International Genomics of Alzheimer’s Project; GWAS, genome-wide association study; MR, Mendelian randomization

### Outcome data

Effect estimates of instrumented pQTLs were extracted from a genome-wide association meta-analysis of clinically diagnosed AD cases within the International Genomics of Alzheimer’s Project (IGAP) [22]. Stage 1 of the GWAS included 21,982 cases and 41,944 controls, all of whom were of European ancestry. Risk of late-onset AD (LOAD) was defined as symptom onset at ≥ 65 years of age. Diagnostic assessment occurred either in clinical evaluation or postmortem autopsy. The mean age at onset for AD cases ranged from 71.1 to 82.6 years, and mean age at examination (or last follow-up) for healthy controls ranged from 51.0 to 78.9 years. Further information on quality control can be found in the original publication.

## Statistical analyses

### Mendelian randomization analysis

MR analyses were conducted to examine the individual effects of each drug target on the risk of AD. Due to the outcome GWAS comprising only autosomal data, we did not include SNPs located within the X chromosome during analysis. To reduce impacts from LD, the instrumented pQTLs underwent a clumping procedure with a threshold of *r*^2^ = 0.001 and a 10,000 kb window to ensure independent variants. The exposure data was coded using the most recent genome reference consortium human build 38 (GRCh38), assembly Hg20, while the IGAP GWAS outcome data used GRCh37 with assembly Hg19. Thus, any instrument pQTLs not found in the outcome dataset were replaced by proxy variants in high LD from the 1000 Genomes European reference panel (*r*^2^ > 0.8). During data harmonization, palindromic SNPs were excluded if the minor allele frequency was > 0.4. MR was performed using the inverse variance weighted (IVW) method. Upon completion of analysis, odds ratios (ORs) and 95% confidence intervals (CIs) for AD outcomes were presented per 1 standard deviation (SD) increase in blood plasma protein concentration.

### Colocalization analysis

Colocalization was performed on *cis*-regions of the encoding target genes of interest (± 500 kb window) to assess if identified associations with AD resulted from shared causal variants. *ACE2* was excluded from this portion of the analysis as it is located on the X chromosome and could not be matched with autosomal outcome data. The following priors were set: SNPs within the default window and exclusively associated with trait 1 had a probability of *p*_1_ = 10^−4^; SNPs within the default window and exclusively associated with trait 2 had a probability of *p*_2_ = 10^−4^; and SNPs within the default window and associated with both traits 1 and 2 had a probability of *p*_12_ = 10^−5^. A posterior probability for shared causal variants (PP.H_4_ ≥ 80%) was considered to demonstrate strong evidence of colocalization. A threshold of PP.H_4_ ≥ 50% was considered suggestive evidence.

## Results

### Effects of DMARD targets on the risk of AD

Overall, there was little evidence in our study that DMARD targets affected the risk of AD (Fig. 3). There was little evidence that the sulfasalazine targets, *PLA2G1B* (OR 0.99; 95% CI [0.91, 1.08]; *p* = 0.80) and *NFKB1* (OR 0.89; 95% CI [0.74, 1.07]; *p* = 0.22), affected AD risk. There was little evidence for the hydroxychloroquine target, *ACE2* (OR 1.10; 95% CI [0.97, 1.24]; *p* = 0.14). There was additionally little evidence for the targets of tocilizumab, the TNF inhibitors, and etanercept, including *LTA* (OR 1.03; 95% CI [0.95, 1.12]; *p* = 0.43), *FCGR2A* (OR 1.02; 95% CI [0.94, 1.10]; *p* = 0.63), *FCGR2B* (OR 0.99; 95% CI [0.87, 1.13]; *p* = 0.86), *TNF* (OR 0.92; 95% CI [0.72, 1.16]; *p* = 0.47), and *IL6R* (OR 0.97; 95% CI [0.90, 1.03]; *p* = 0.28). However, there was evidence that increasing plasma levels of *FCGR3B*, an etanercept target, increased the risk of AD (OR 1.10; 95% CI [1.02, 1.19]; *p* = 0.01).

**Fig. 3.**
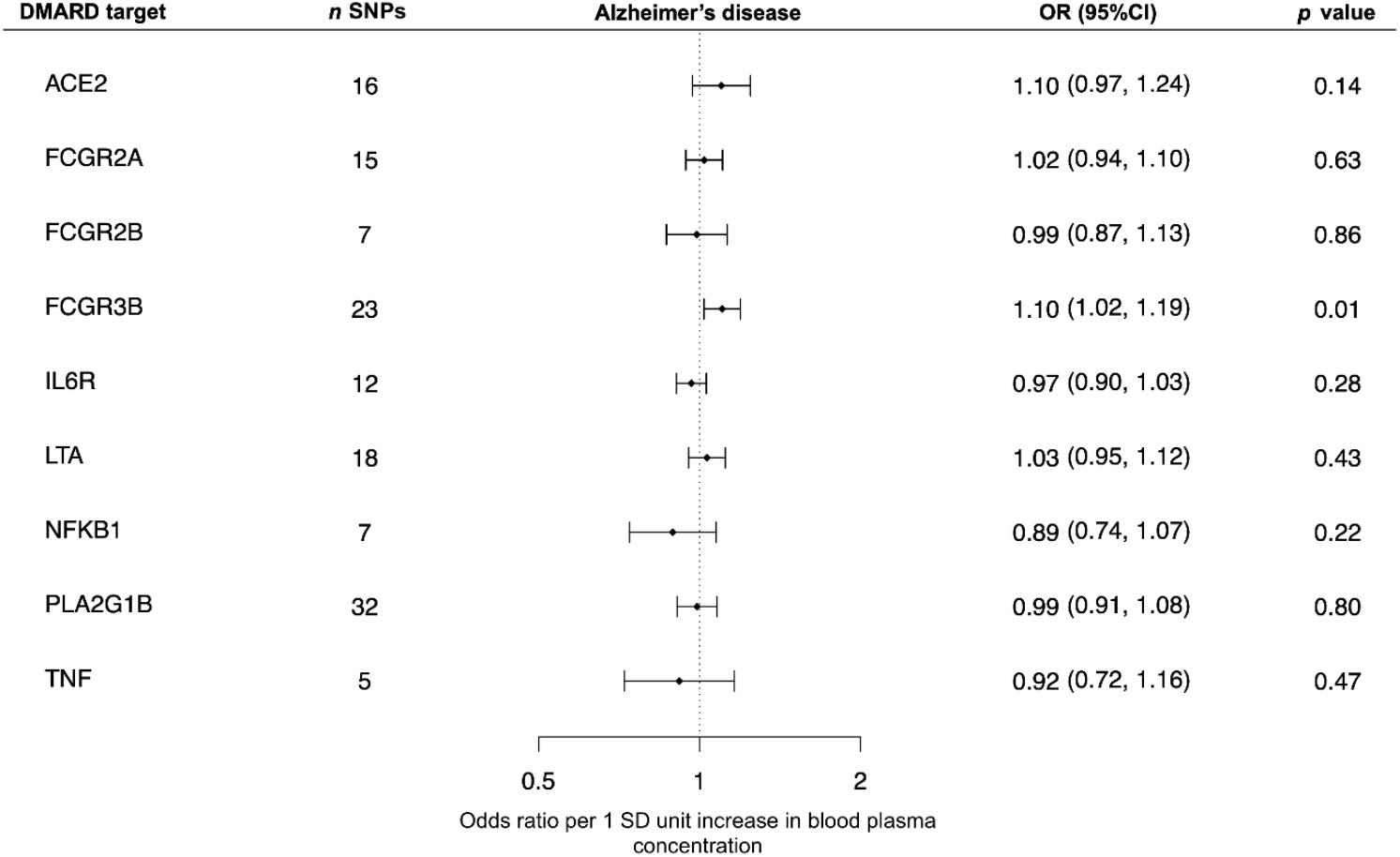
Estimates of the effects of selected DMARD targets on the risk of developing AD. Error bars correspond to the 95% confidence intervals. Abbreviations: DMARD, disease-modifying antirheumatic drug; SNP, single nucleotide polymorphism; OR, odds ratio; SD, standard deviation

### Results of colocalization analysis

Colocalization analyses were consistent with the MR analyses above. There was little evidence of colocalization between any of the targets and AD (PP.H_4_ < 10% across all proteins). Despite evidence of a causal effect in the main MR analysis, *FCGR3B* protein and AD did not colocalize in the flanked chromosome 1 region of *FCGR3B* (PP.H_4_ = 6.21%). There was also very little evidence of pleiotropy (PP.H_3_ = 10.3%). However, there was some evidence of a non-colocalized association between *TNF* protein and AD, potentially through pleiotropic causal variants (PP.H_3_ = 46.5%) (Fig. 4).

**Fig. 4.**
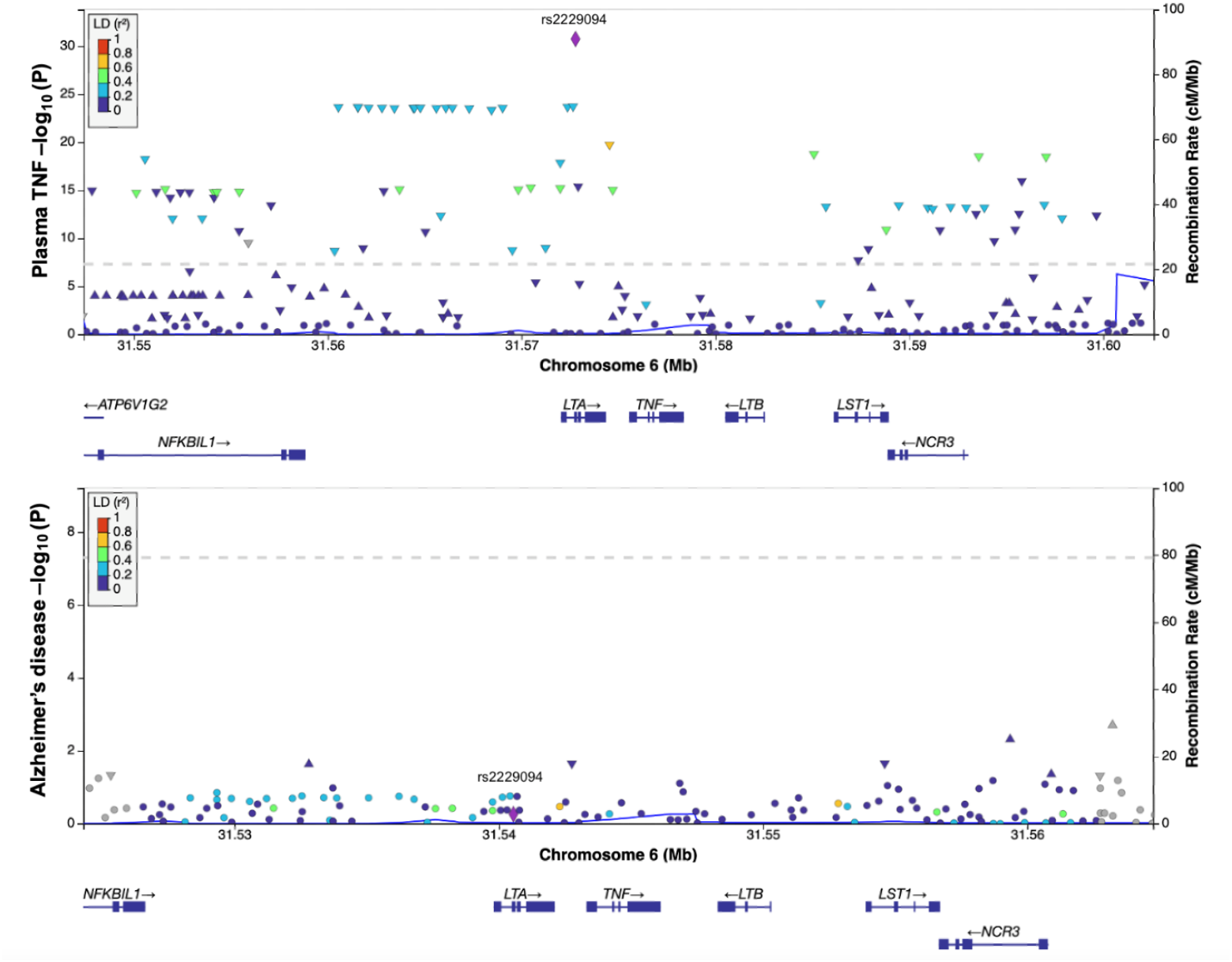
Regional genomic plots from colocalization analysis of the flanked *TNF* region on chromosome 6. The LD reference SNP (rs2229094) was used for plotting. Both traits (*TNF*, AD) had to be compared separately because of the differing builds, GrCH37/38. No pQTLs in the *TNF* region met the threshold for colocalization with AD.

## Discussion

We utilized *cis*-pQTL and GWAS summary statistics to evaluate the causal effects of genetically perturbed DMARD targets on the risk of AD. To our knowledge, this is the first study to specifically examine DMARD targets and AD risk using drug target MR and colocalization.

We found limited evidence to support the repurposing potential of select DMARD targets for reduced AD risk. The MR analyses provided little evidence that 8 out of the 9 selected targets— *ACE2, FCGR2A, FCGR2B, IL6R, LTA, NFKB1, PLA2G1B*, and *TNF*—affected the risk of AD. With MR, we found that a per 1 SD unit increase in plasma *FCGR3B* concentration leads to a 10% greater risk of AD. Though, further analysis of genetic signaling in the flanked region on chromosome 1 revealed that *FCGR3B* did not colocalize with AD and are thus unlikely to share a single causal variant. A possible explanation for this discordance is that the findings from MR may have been driven by distinct variants in LD. Alternatively, the lack of evidence for colocalization may be due to tissue specificity. In support of this, a recent study found evidence of colocalization between *FCGR3B* protein, located in cerebrospinal fluid (CSF), and AD signals [23]. Interestingly, this study observed the opposite direction of effect for *FCGR3B*, with their MR analysis revealing an association between increased CSF *FCGR3B* concentration and a lower risk of AD—although it should be noted that their instruments for *FCGR3B* did not reach genome-wide significance. CSF protein composition is more diverse than that of plasma and may perform as a more accurate biomarker for neurodegenerative diseases [24]. However, plasma, while sub-optimal in this case, has greater availability of proteomic data, rendering it a useful and more accessible tool for analysis. It is possible that the use of a less disease-relevant tissue such as blood plasma may explain the difference in results, though, both sets of findings are indicative of causal effects of *FCGR3B* on the risk of AD.

The evidence in our study suggests that *ACE2, LTA, FCGR2A, FCGR2B, and PLA2G1B* are unlikely to be effective candidate targets for AD; these findings should be taken into consideration to reduce the high attrition rate in AD drug development by discouraging trials of irrelevant targets such as those listed above [25]. Additionally, we observed little evidence of an effect for *IL6R*, though our estimate shows a consistent direction of effect with that of a recent MR study reporting significant protective effects of *IL6R* variants on AD risk via downstream C-reactive protein serum levels [26]. As that study utilized data from larger-scale GWASs, including ‘proxy’ AD cases—as opposed to the present study which only examined clinically diagnosed AD cases—it is possible that with greater power, we may have similarly observed a significant effect of *IL6R*. Alternatively, it is plausible that they observed an effect of *IL6R* due to bias introduced by the inclusion of ‘proxy’ AD cases. Definitive conclusions cannot be drawn regarding the efficacy of *NFKB1* and *TNF* due to the imprecision of estimates observed in this study. However, a previously conducted MR found little evidence of a causal effect of *TNF* serum levels on AD [27]. Their instrumentation of pQTLs derived from blood serum raises the concern of tissue relevancy as mentioned earlier in the discussion of *FCGR3B*. Our suggestive evidence of a non-colocalized association between *TNF* and AD signals at the flanked chromosome 6 region highlights the possibility of alternate tissue-specific molecular pathways in which *TNF* and AD associate. Nevertheless, a potential ∼28% reduction in disease risk through *TNF* plasma levels, as evidenced by this study, merits further exploration of this target in plasma tissue.

## Limitations

Several limitations should be considered. First, this study was limited by the lack of proteomic data available for relevant drug targets. Among 33 initially selected targets from a total of 11 DMARDs, the final MR analyses could only include 9 targets from 8 DMARDs. Notably, methotrexate had to be excluded, which has recently accumulated promising observational evidence for an association with reduced AD risk [28]. Second, clinical evidence has demonstrated differential protein expression according to tissue specificity and it is plausible that this would have impacted our findings, in addition to the sub-optimal relevance of plasma as a biomarker for neurodegenerative processes [29]. Additional analyses that incorporate locus-specific instruments in tissues other than plasma (e.g., CSF) may yield different results. Third, there are several caveats to drug target MR that limit the ability to causally examine broader drug usage and disease outcome associations [30]. As we estimated the effects of perturbing individual drug targets, we cannot infer the likely aggregate effects of specific DMARDs. Furthermore, it is difficult to achieve a complete understanding of the pharmacodynamics of medications more broadly (e.g., understanding the exact molecular pathways in which etanercept affects *FCGR3B* and other targets). Thus, based on the genetic evidence alone, we cannot conclude how prescribed etanercept usage would affect AD risk via *FCGR3B* and other known or unknown targets. Further mechanistic studies and clinical trials are recommended for comprehensive evaluation of DMARD repurposing. Finally, it is worth noting possible biases which could have further attenuated our estimates. Sourcing instruments from the UK BioBank—which is prone to selective representation—may introduce collider bias [31, 32]. Selected study participants tend to be healthier and may therefore have a lower genetic liability for disease traits, which in turn may potentially impact MR estimates [33]. That said, to date, there is little evidence in the literature specifically linking proteomic data to selection bias.

## Conclusions

The findings from our study do not support the majority of selected DMARD targets as efficacious treatment candidates for AD, with exception of the etanercept target, *FCGR3B*. The observed positive effect of increased plasma *FCGR3B* concentration on AD risk warrants further investigation into the viability of therapeutics which lower *FCGR3B* protein levels in blood plasma. Our findings should be considered as part of the current discourse regarding the targeting of inflammatory mechanisms for AD treatment and prevention. Larger-scale MR studies that instrument a greater number of DMARD targets across various tissues are additionally recommended.

## Data Availability

All data produced in the present work are contained in the manuscript and available at

https://github.com/tkush15/AD_DMARD_MRCOLOC

## Abbreviations

AD: Alzheimer’s disease
MR: Mendelian randomization
DMARD: Disease-modifying antirheumatic drug
pQTL: Protein quantitative trait loci
CNS: Central nervous system
TNF-α: Tumor necrosis factor
IL-6: Interleukin-6
GWAS: Genome-wide association study
RCT: Randomized control trial
LD: Linkage disequilibrium
BMI: Body mass index
UKB-PPP: UK Biobank Pharma Proteomics Project
SNP: Single nucleotide polymorphism
IGAP: International Genomics of Alzheimer’s Project
LOAD: Late-onset Alzheimer’s disease
GRCh: Genome Reference Consortium Human
IVW: Inverse variance weighted
OR: Odds ratio
CI: Confidence interval
SD: Standard deviation

## Declarations

### Ethics approval and consent to participate

The GWASs included in this study obtained written informed consent from all participants prior to data collection. Related study protocols were approved accordingly by the appropriate institutional review boards accordingly. Therefore, separate ethical approval was not required for the present study.

### Consent for publication

Not applicable.

### Availability of data and materials

Proteogenomic summary data from the UKB-PPP can be found at http://ukb-ppp.gwas.eu. Full AD GWAS data can be obtained by requesting access via the National Institute on Aging Genetics of Alzheimer’s Disease Data Storage Site (NIAGADS) data repository, ID: NG00075. The codes used to generate the results of this study are located at https://github.com/tkush15/AD_DMARD_MRCOLOC.

### Authors’ contributions

NMD, ELA, and CNK conceptualized the study. ELA provided coding. VTB provided coding and guidance for colocalization. CNK performed all statistical analyses, prepared figures, and drafted the initial manuscript. NMD and ELA guided the interpretation of results and provided written edits to the manuscript draft. All authors contributed to the revision and approval of the final manuscript.

## Acknowledgments

Not applicable.

